# The prevalence of antibodies to SARS-CoV-2 among blood donors in China

**DOI:** 10.1101/2020.07.13.20153106

**Authors:** Le Chang, Wangheng Hou, Lei Zhao, Yali Zhang, Yanbin Wang, Linfeng Wu, Tingting Xu, Lilin Wang, Juan Wang, Jian Ma, Lan Wang, Junpeng Zhao, Jing Xu, Juan Dong, Ying Yan, Ru Yang, Yu Li, Fei Guo, Wenjuan Cheng, Yingying Su, Jinfeng Zeng, Wei Han, Tong Cheng, Jun Zhang, Quan Yuan, Ningshao Xia, Lunan Wang

**Author notes:** These authors contributed equally to this article (Le Chang, Wangheng Hou, Lei Zhao, Yali Zhang). Corresponding author: Lunan Wang, Address: No.1 Dahua Road, Beijing 100730, P.R. China, or Quan Yuan, Address: No. 4221 Xiang’an South Road, Xiamen 361102, P.R. China.

## Abstract

**Objectives:** The prevalence of antibodies to SARS-CoV-2 among blood donors in China remains unknown. To reveal the missing information, we investigated the seroprevalence of SARS-CoV-2 antibodies among blood donors in the cities of Wuhan, Shenzhen, and Shijiazhuang of China.

**Design:** Cross-sectional study

**Setting:** Three blood centers, located in the central, south and north China, respectively, recruiting from January to April 2020.

**Participants:** 38,144 healthy blood donors donated in Wuhan, Shenzhen and Shijiazhuang were enrolled, who were all met the criteria for blood donation during the COVID-19 pandemic in China.

**Main outcome measures:** Specific antibodies against SARS-CoV-2 including total antibody (TAb), IgG antibody against receptor-binding domain of spike protein (IgG-RBD) and nucleoprotein (IgG-N), and IgM. Pseudotype lentivirus-based neutralization test was performed on all TAb-positive samples. In addition, anonymous personal demographic information, including gender, age, ethnicity, occupation and educational level, and blood type were collected.

**Results:** A total of 519 samples from 410 donors were confirmed by neutralization tests. The SARS-CoV-2 seroprevalence among blood donors was 2.29% (407/17,794, 95%CI: 2.08% to 2.52%) in Wuhan, 0.029% (2/6,810, 95%CI: 0.0081% to 0.11%) in Shenzhen, and 0.0074% (1/13,540, 95%CI: 0.0013% to 0.042%) in Shijiazhuang, respectively. The earliest emergence of SARS-CoV-2 seropositivity in blood donors was identified on January 20, 2020 in Wuhan. The weekly prevalence of SARS-CoV-2 antibodies in Wuhan’s blood donors changed dynamically and were 0.08% (95%CI: 0.02% to 0.28%) during January 15 to 22 (before city lockdown), 3.08% (95%CI: 2.67% to 3.55%) during January 23 to April 7 (city quarantine period) and 2.33% (95%CI: 2.06% to 2.63%) during April 8 to 30 (after lockdown easing). Female and older-age were identified to be independent risk factors for SARS-CoV-2 seropositivity among donors in Wuhan.

**Conclusions:** The prevalence of antibodies to SARS-CoV-2 among blood donors in China was low, even in Wuhan city. According to our data, the earliest emergence of SARS-CoV-2 in Wuhan’s donors should not earlier than January, 2020. As most of the population of China remained uninfected during the early wave of COVID-19 pandemic, effective public health measures are still certainly required to block viral spread before a vaccine is widely available.

## Introduction

Coronavirus disease 2019 (COVID-19), first found in December 2019, fast spread all over the world and was declared as a pandemic in March 2020.^1^ The total of confirmed cases has surpassed 12 million, involving 216 countries, and more than 566,000 people died resulting from acute respiratory diseases and its related complications.^2^ A novel beta-coronavirus was identified as the original etiological agent of COVID-19. The genome of the new virus was 70% similar to that of severe acute respiratory syndrome coronavirus (SARS-CoV), and it was designated SARS-CoV-2.^3,4^

Nucleic acid testing of SARS-CoV-2 from the upper or lower respiratory tract, feces, urine, or other specimens could quickly identify infected people from suspected cases. The timely diagnosis could help reduce patient gathering and shorten the length of stay in clinics, thus promoting effective infection control management.^5^ However, the COVID-19 pandemic is like an iceberg:^6^ what we can see is those who are severe cases, part of mild to moderate cases, and known asymptomatic cases diagnosed by the screening of close contacts of COVID-19 cases or random testing of a specific population. An uncertain number of asymptomatic individuals and parts of mild cases may be missed, like the bottom of the iceberg under the water. These asymptomatic individuals may contribute to the transmission of the disease.^7,8^ Moreover, it is crucial to identify the asymptomatic infections to estimate the disease burden and to get a better understanding of the real case fatality rate.^9,10^ The missing information could be obtained by screening the population for specific antibodies using validated serologic assays.

Host humoral immune response to SARS-CoV-2 among COVID-19 confirmed patients had been characterized by several studies.^11-14^ Total antibody (TAb) specific to SARS-CoV-2, which had been demonstrated to be the most sensitive and earliest serologic biomarker, usually increasing since the second week of symptoms onset,^12^ and two weeks after onset, all infected cases showed reactive results.^11^ In contrast, the IgM and IgG seroconversion generally occurred on the second or third week,^13^ following a quick decrease of IgM and a longtime IgG persistence.^12^ So far, only limited information on serologic screening of specific asymptomatic people showed that seroprevalence of SARS-CoV-2 varied from 1.6% to 4.1% among different countries and populations.^15-17^ However, comprehensive data of antibody response against SARS-CoV-2 in asymptomatic individuals of mainland China are unclear.

In this study, we investigated the prevalence of SARS-CoV-2 antibodies among donors who donated their blood from January to April 2020 in the cities of Wuhan, Shenzhen, and Shijiazhuang.

These three different cities, locating in the central, south and north of China, have similar population size but of distinct COVID-19 incidence. Moreover, the potential risk factors for SARS-CoV-2 seropositivity were analyzed.

## Methods and Materials

### Study design and participants

Blood donors donated from January to April in Wuhan, Shenzhen, and Shijiazhuang were enrolled in the study. The population size, number of confirmed COVID-19 cases, and donation time of enrolled blood donors in three different cities were listed in Table 1. Criteria used for all blood donors during the pandemic are listed as enclosed: 1) have neither a fever (body temperature ≤37.3°C) nor any respiratory symptoms for at least 28 days; 2) have neither close contact to those confirmed or suspected COVID-19 cases or clustering occurrence of cases for at least 28 days; 3) Body temperature is normal before donation (≤37.3°C); 4) in Shenzhen and Shijiazhuang, donors who have a history of residence in or travel from Hubei province or have close contact to people from Hubei are suggested to defer blood donation for at least 28 days. A total of 38,144 blood donors were enrolled in this study. Anonymous personal demographic information, including gender, age, ethnicity, occupation and educational level, and blood type were collected.

**Table 1.** The population size, number of confirmed COVID-10 cases and donation time of enrolled blood donors in three different cities.

| Blood Center | City | Population*<br>(million) | Number of<br>confirmed COVID-19 cases **<br>(prevalence<br>/100,000) | Donation time of<br>enrolled blood<br>donors |
| --- | --- | --- | --- | --- |
| Wuhan Blood<br>Center | Wuhan | 11.21 | 50,333 (448.92) | 15 Jan - 30 Apr |
| Shenzhen Blood<br>Center | Shenzhen | 13.03 | 420 (3.22) | 2 Feb - 15 Mar |
| Hebei Province<br>Blood Center | Shijiazhuang | 11.03 | 29 (0.26) | 23 Jan - 15 Mar |

\*The population data was collected from the statistics released by the Hubei Provincial, Shenzhen, and Hebei Provincial Bureau of statistics in 2019.
\*\*Data were collected from the website of the Hubei Province Health Committee, Shenzhen Health Committee, and Hebei Province Health Committee.

### Serological tests

After routinely tested for transfusion-transmitted pathogens (including HIV antibody), all donation samples were tested for total antibody (TAb) against SARS-CoV-2, and the reactive samples were further tested for SARS-CoV-2 specific IgG and IgM antibodies. All the serologic screening tests used enzyme-linked immunosorbent assay (ELISA) kits provided by Beijing Wantai Biological Pharmacy Enterprise Co.,Ltd. The detection experiments were performed according to the manufacturer’s instructions. In brief, TAb detection was based on a double-antigens sandwich immunoassay, using two kinds of mammalian cell-expressed recombinant antigens contained the receptor-binding domain (RBD) of the spike protein of SARS-CoV-2 as the immobilized and HRP-conjugated antigen, respectively. Two kinds of specific IgG antibodies were tested using an indirect ELISA method based on recombinant antigens, RBD antigen (IgG-RBD), and nucleoprotein (IgG-N), respectively. The IgM μ-chain capture method was used to detect the IgM antibodies, using the same HRP-conjugate RBD antigen. Those IgG or IgM positive samples with the signal to the cutoff ratio (S/CO) ≥10 were further diluted (1:10, 1:40, 1:160…, and 1:40960) by normal saline and tested again. The titer for IgG and IgM antibodies was calculated via S/CO multiplied by the maximum dilution factor.

### Pseudotype lentivirus-based neutralization test (ppNAT)

For confirmation of the presence of neutralizing antibodies, all the TAb positive donation samples were tested against lentiviral pseudotyping particle (LVpp) bearing SARS-CoV-2 spike antigen. The production and detailed information regarding the assay were described in Supplementary Material. All samples were tested in serial dilutions following procedure illustrated in Supplementary Material (Figure S1), and the pseudotyping particle-based neutralization titer (ppNAT) of each sample, which is defined as the maximum dilution fold required to achieve infection inhibition by 50% (ID50), was determined by the 4-parameter logistic (4PL) regression. An ID50 ≥20 was determined as a cutoff value for the presence of neutralizing antibodies.

### Statistical analysis

The binary logistic regression was used to investigate the role of individual demographic factors on the outcome of specific antibodies against SARS-CoV-2: single-factor analysis was first used to discriminate variables with a statistical difference, then multivariate regression analysis was performed to adjust the odds ratio (OR) of these variables. Kruskal-Wallis test or Mann-Whitney U were used to compare more than two or two groups of independent data, respectively. All these statistical analyses above were realized by SPSS v21.0 (IBM SPSS, Chicago, IL). Linear correlation analysis between different antibodies and ID50 was calculated by GraphPad Prism v8.0 (GraphPad Software, San Diego, CA) or Origin 2020 (OriginLab, Northampton, MA). p<0.05 was considered statistically significant.

### Patient and public involvement

This was a retrospective study and no patients or public were directly involved in the study design, setting the research questions, or the outcome measures. No patients or public were asked to advise on the interpretation of the results.

## Results

### Characteristics of enrolled blood donors

Almost all blood donors donated in the three blood centers during the study period were enrolled, totaling 38,144 blood donors, including 17,794 were from Wuhan, 6810 from Shenzhen, and 13,540 from Shijiazhuang. The characteristics of the involved donors were summarized in Table 2. The median age was 33 (IQR, 19 to 47), 36 (IQR, 19 to 53), and 40 (IQR, 33 to 48) for donors from the three cities, respectively. Among these donors, 29.5% to 37.7% were female. Moreover, 2329 donors donated twice or more during January to April 2020, including 1559 in Wuhan, 221 in Shenzhen, and 549 in Shijiazhuang.

**Table 2.**
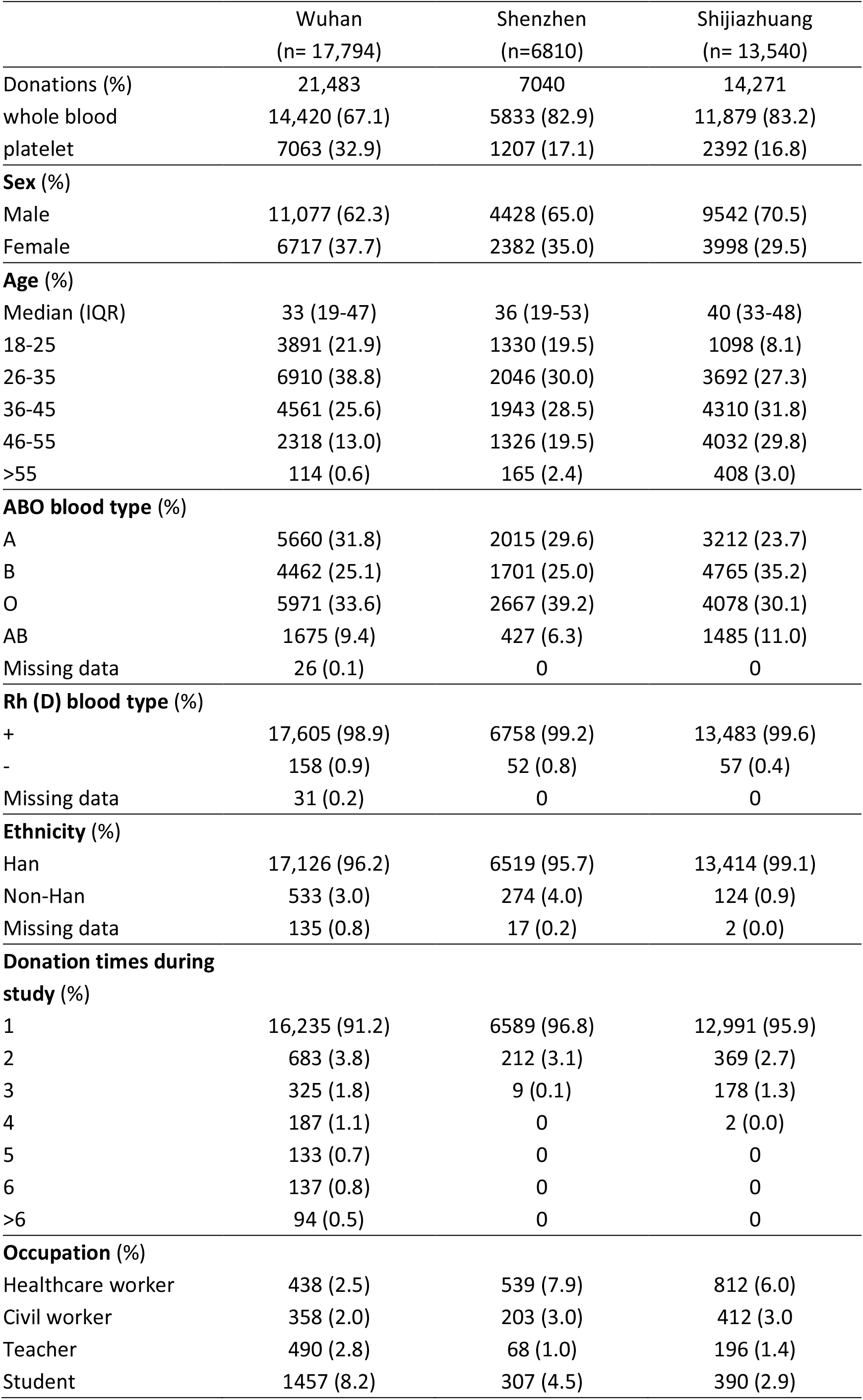
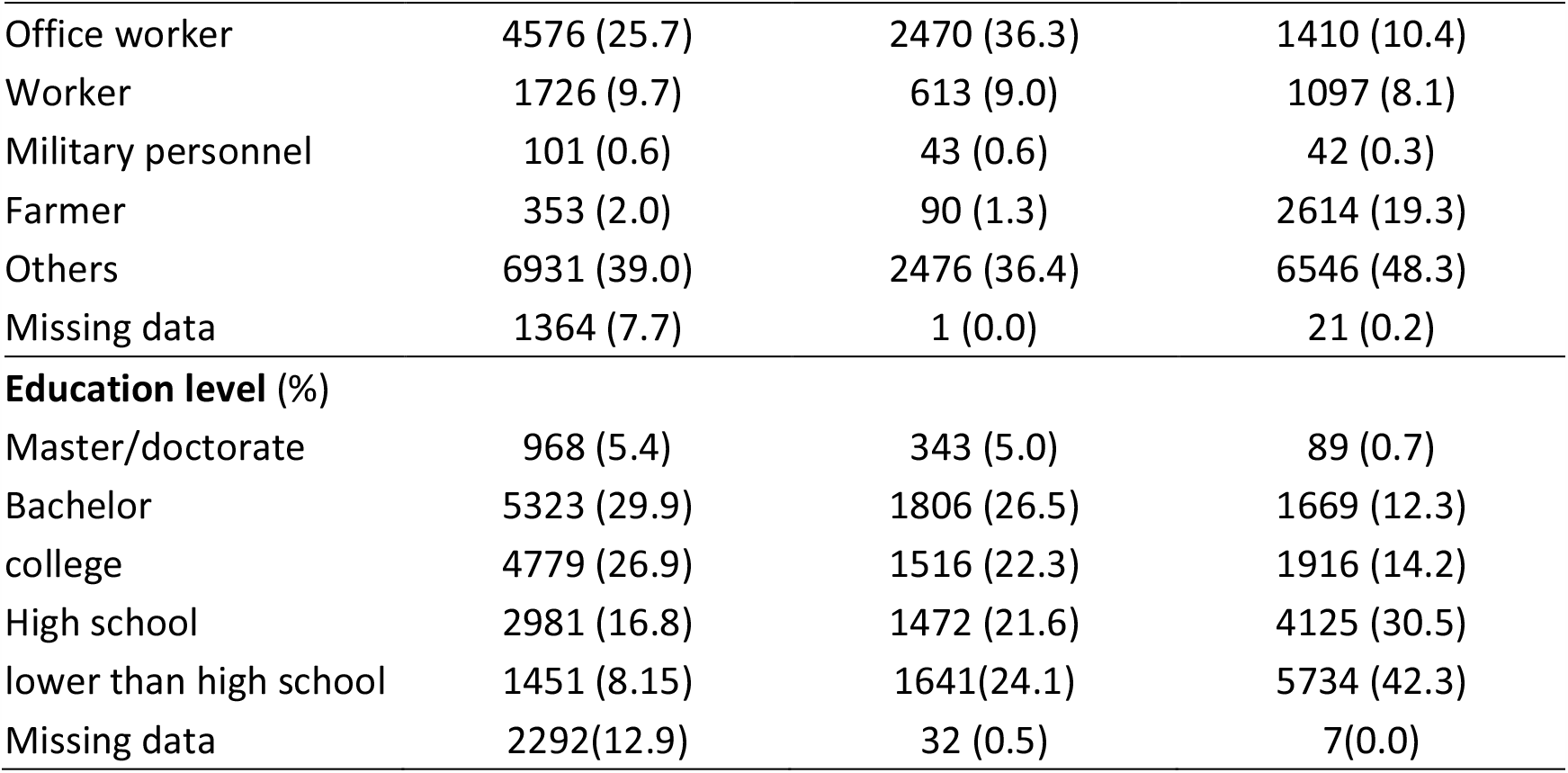
Characteristics of enrolled blood donors

### Antibody prevalence of SARS-CoV-2 among blood donors

A total of 42,794 blood donation samples were tested for specific total antibody against SARS-CoV-2 in the study, including 32,282 whole blood and 10,712 platelet donations. Among all samples, 678 (678/42,794, 1.58%) donations were TAb positive (all were HIV-free), of which 590 (590/21,483, 2.75%) were from Wuhan, 28 (28/7040, 0.40%) from Shenzhen, and 60 (60/14,271, 0.42%) from Shijiazhuang. All the TAb positive samples were further tested for SARS-CoV-2 IgG-RBD, IgG-N, IgM and ppNAT. Finally, 519 donation samples from 410 donors were confirmed the presence of SARS-CoV-2 neutralization antibody by the ppNAT tests. The screening and confirmatory procedures were shown in Figure 1. The results revealed that the SARS-CoV-2 seroprevalence in the three different populations were 2.29% (407/17,794, 95%CI: 2.08% to 2.52%) in Wuhan, 0.029% (2/6,810, 95%CI: 0.0081% to 0.11%) in Shenzhen, and 0.0074% (1/13,540, 95%CI: 0.0013% to 0.042%) in Shijiazhuang, respectively. Among all confirmed-seropositive donations, 96.34% (500/519) were IgG-RBD positive, 78.42% (407/519) were IgG-N positive, and IgM antibody was detectable in 343 (66.09%, 343/519) donations.

**Figure 1.**
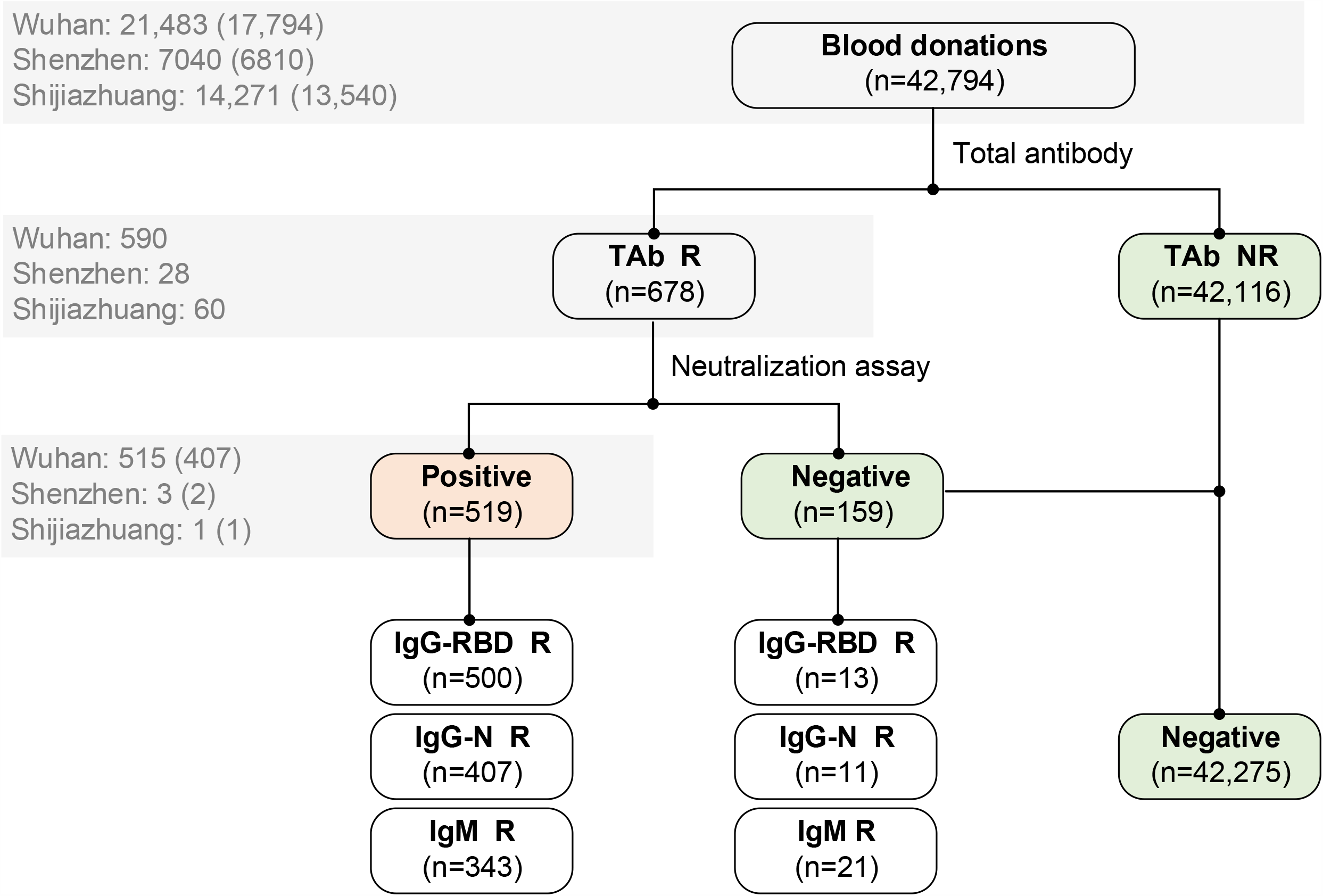
Flow chart of screening and confirmatory procedure. A total of 42,794 blood donations were tested for total antibody. Those reactive were further confirmed by neutralization assay and tested for IgG-RBD, IgG-N, and IgM antibodies against SARS-CoV-2. Five hundred nineteen donations from 410 donors were finally confirmed. Numbers of donations (blood donors) tested for specific antibodies in three cities are shown in the grey box. The seroprevalence of antibodies to SARS-CoV-2 among blood donors was calculated by the number of confirmed positive donors divided by the total number of tested donors: 2.29% (407/17,794, 95%CI: 2.08%-2.52%) in Wuhan, 0.029% (2/6810, 0.0081%-0.11% in Shenzhen, and 0.0074% (1/13,540, 95%CI: 0.0013%-0.042%) in Shijiazhuang, respectively. R, reactive; NR, non-reactive; IgG-RBD, IgG antibody against receptor-binding domain (RBD) of the spike protein of SARS-CoV-2; IgG-N, IgG antibody against nucleoprotein of SARS-CoV-2.

In samples from Wuhan involved in our study, there were 2607 blood donations that were collected before January 23, 2020 when Wuhan was quarantined, including 1614 samples were donated during January 15 to 18 (week 3 of 2020) and 993 samples were donated during January 19 to 22. Among these donations, two were confirmed for SARS-CoV-2 seropositivity. The data suggested that the SARS-CoV-2 seroprevalence in the blood donor population of Wuhan from mid to late January 2020 was about 0.08% (2/2607, 95%CI: 0.02% to 0.28%). During the Wuhan quarantine period (January 23 to April 7), 256 donations with confirmed serological evidence from 185 donors were identified from 7903 samples of 6004 Wuhan’s blood donors, suggesting a seroprevalence of 3.08% (185/6004, 95%CI: 2.67% to 3.55%). After April 8 when the city was eased (April 8 to 30), we further tested a total of 10,973 samples of 10,708 donors, and found out that 257 samples of 249 donors were confirmed SARS-CoV-2 seropositive. The SARS-CoV-2 seroprevalence among Wuhan’s blood donors during this period was 2.33% (249/10708, 95%CI: 2.06% to 2.63%). In a time series analysis (Figure 2, upper panel), the highest seroprevalence (4.63%, 12/259, 95%CI: 2.67% to 7.92%) was observed among the Wuhan’s blood donors at 6^th^ week of 2020 (February 2 to 8), about 3-week after the quarantine. Since then, the seropositive rate showed a clearly downward trend in blood donors of Wuhan. In contrast, the positive rates were very low in samples from donors of Shenzhen and Shijiazhuang, which were all collected during the Wuhan quarantine period (Figure 2, middle and lower panels).

**Figure 2.**
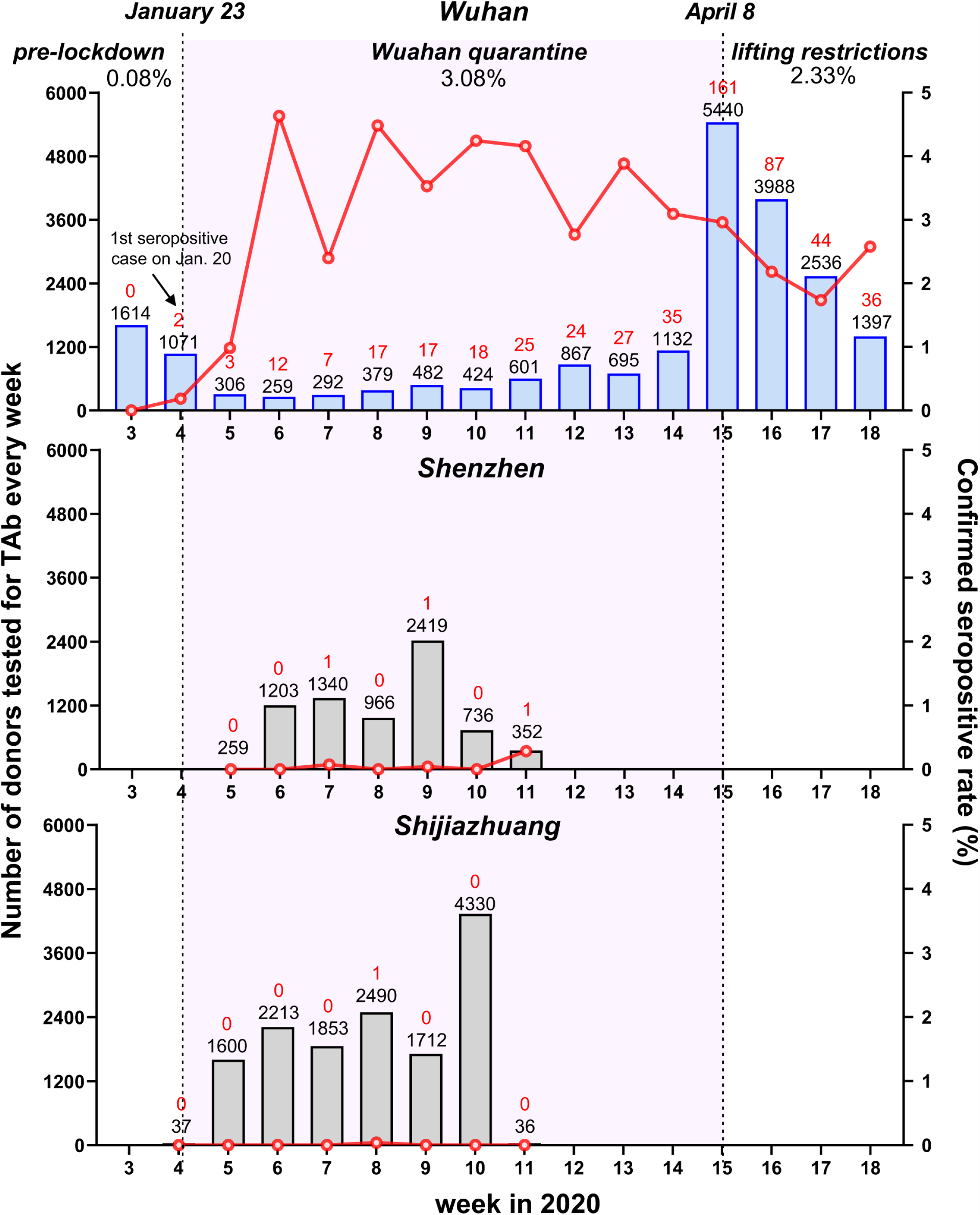
Weekly seroprevalence of SARS-CoV-2 antibody during different periods from January to April 2020 in the cities of Wuhan, Shenzhen and Shijiazhuang. The number of donors tested for total antibody (TAb) every week (the black numbers on the top of each histogram) is shown in histograms. The number of confirmed positive cases is shown in red numbers on the top of each histogram. The confirmed seropositive rate (number of ppNAT-confirmed donors/number of donors tested for TAb) in each week is shown in red lines. The first donor confirmed positive by the pseudotype lentivirus-based neutralization tests in Wuhan were on January 20, the fourth week of 2020. Lockdown of Wuhan City started on January 23 and on April 8, all the travel restrictions in Wuhan were lifted. The period of study in Wuhan is divided into three stages: pre-lockdown (Jan 15-Jan 22), lockdown (Jan 23-Apr 7), and lifting restrictions (Apr 8-Apr30). The confirmed seroprevalences of the three stages varied: two from 2607 donors were confirmed in the first stage (0.08%, 95%CI: 0.02%-0.28%); 256 donations with confirmed serological evidence from 185 donors were identified from 7903 samples of 6004 donors, suggesting a seroprevalence of 3.08% (95%CI: 2.67%-3.55%) in the lockdown stage; After April 8, we further tested a total of 10,973 samples of 10,708 donors, and found out that 257 samples of 249 donors were confirmed SARS-CoV-2 seropositive (2.33%, 95%CI: 2.06%-2.63%). The peak of seroprevalence (4.63%, 12/259) occurred in the stage of lockdown.

### Associations between the titer of ppNAT and other antibody markers

We analyzed the associations between the titer of ppNAT (ID50) with TAb (undiluted S/CO), IgG-RBD (dilution quantitative S/CO), IgG-N (dilution quantitative S/CO) and IgM (dilution quantitative S/CO) among all 678 donations with detectable TAb. As the results shown in Figure 3A, the average ppNAT titer successively elevated with the increasing of TAb S/CO value. Notably, the percentage of samples with a ppNAT ID50 ≥20 (confirmatory presence of neutralization antibody) were 38.07% (83/218), 80.65% (75/93), 91.67% (66/72) and 98.99% (295/298) among those with TAb S/CO strata of 1-5, 5-10, 10-15, and >15, respectively. Moreover, the ppNAT titer was positively correlated with the titers of IgG-RBD (r=0.823, p<0.001, Figure 3B), IgG-N (r=0.725, p<0.001, Figure 3C) and IgM (r=0.617, p<0.001, Figure 3D), respectively. Overall, the IgG-RBD titer showed the best correlation with the confirmatory ppNAT titer among TAb-positive samples. The average titer of IgG-N was significantly lower than that of IgG-RBD (p<0.001), suggesting the antibody response against viral nucleocapsid (N) protein may be weaker than that to viral spike protein.

**Figure 3.**
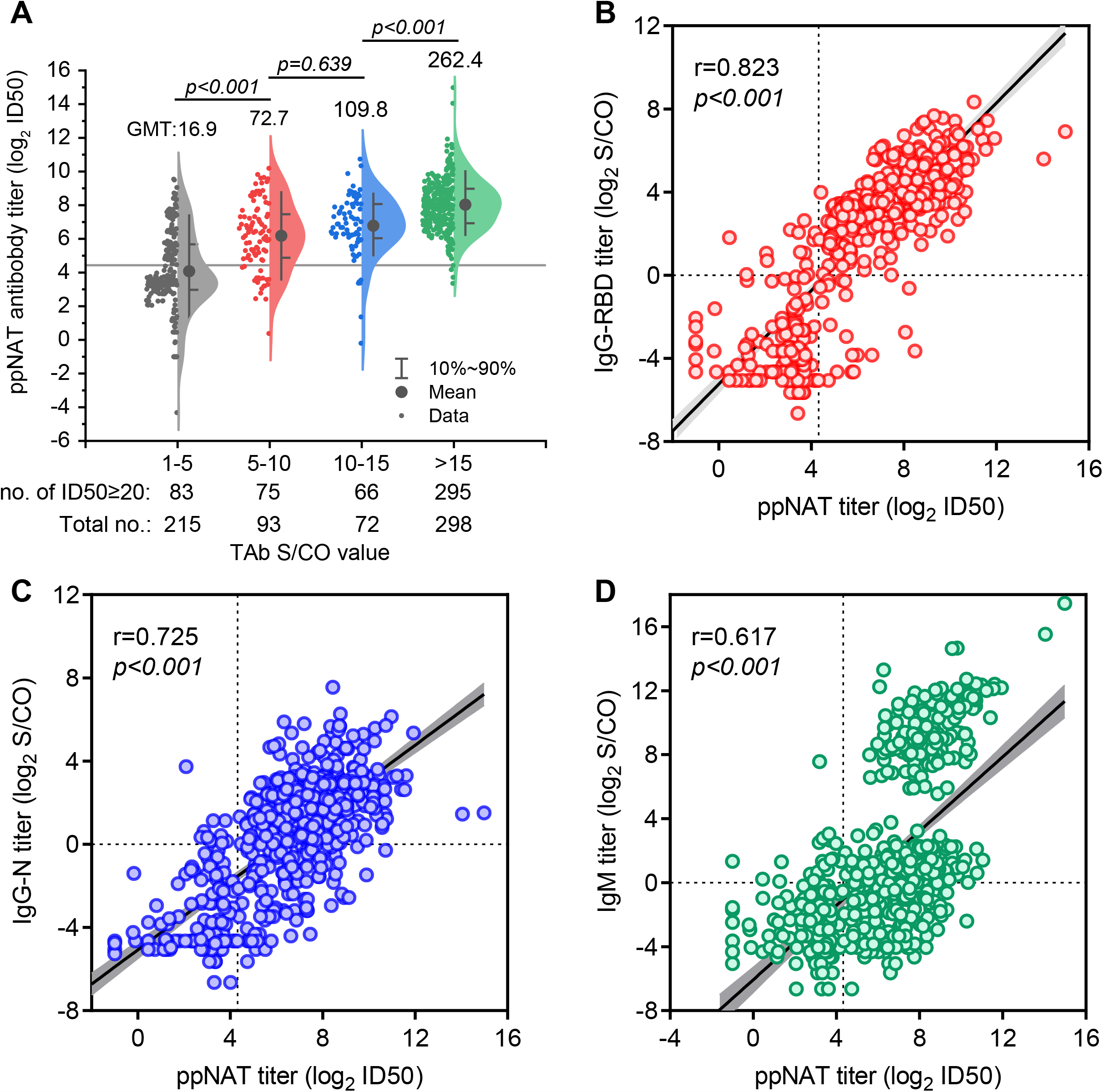
Relationships between ppNAT titer and specific antibodies titer among all 678 TAb-positive samples A. The relationship between ID50 of pseudotype lentivirus-based neutralization tests (ppNAT) and different groups of signal to the cutoff ratio (S/CO) of total antibody. The horizontal grey line shows the cutoff value of ppNAT (ID50=20). The neutralization titer successively elevated with increasing S/CO of TAb among all samples (p<0.001). Group 1-5 vs. 5-10: p <0.001; Group 1-5 vs. 10-15: p <0.001; Group 1-5 vs. 15-20: p <0.001; Group 5-10 vs. 10-15: p =0.639; Group 5-10 vs. 15-20: p <0.001; Group 10-15 vs. 15-20: p <0.001. Geometric mean titers (GMT) of the four groups are 16.9, 72.7, 109.8, and 262.4, respectively. B. The relationship between ppNAT ID50 titer and titer of IgG-RBD antibody. The correlation coefficient (r) by linear correlation analysis between IgG-RBD and ID50 was as high as 0.823 (p<0.001). The dotted lines show the cutoff value of ppNAT (ID50=20) and IgG-RBD assay (S/CO=1.0). C. The relationship between ppNAT ID50 titer and titer of IgG-N antibody. The correlation coefficient (r) between IgG-N and ID50 was 0.725 (p <0.001). The dotted lines show the cutoff value of ppNAT (ID50=20) and IgG-N assay (S/CO=1.0). D. The relationship between ppNAT ID50 titer and titer of IgM antibody. The correlation coefficient (r) between IgM and ID50 is 0.617 (p<0.001). The dotted lines show the cutoff value of ppNAT (ID50=20) and IgM assay (S/CO=1.0).

Although the high sensitivity and specificity of the double-sandwich RBD-based TAb ELISA had been demonstrated in COVID-19 patients and asymptomatic infections, the positive predictive value (PPV) of this test was significantly varied among different populations when it was used alone. In this study, the PPV of TAb ELISA was 87.29% (515/590) in samples from Wuhan, 10.71% (3/28) in samples from Shenzhen, and was 1.67% (1/60) in samples from Shijiazhuang. Overall, the PPV values of TAb in different populations were highly-associated with the reported numbers of COVID-19 confirmed cases of these cities. These results highlighted the necessity of the confirmatory testing for TAb-positive samples, in particular for those from the non-epidemic area. To establish an optimized strategy with practical applicability for the situation when a cell-based neutralization test is unavailable, we analyzed the diagnostic performance for combined use of TAb and other immunoassay-determined antibody markers. Table 3 listed the added diagnosis value of IgG-RBD, IgG-N, and IgM in TAb-positive samples from Wuhan (n=590) and the other two cities (n=88). Apparently, additional IgG-RBD tests in TAb-positive samples was a preferred screening approach, which showed a significantly improved PPV (TAb/IgG-RBD double reactive, PPV: 99.01%) with little missing (17/515, 2.33%) of true-positive samples among blood donors in Wuhan (Table 3). However, this combined strategy showed minimal PPV improvement among donors in Shenzhen and Shijiazhuang (Table 3)

**Table 3.** Comparison of performance of the combined markers in the detection of confirmatory SARS-CoV-2 antibody among blood donors in Wuhan and other cities.

| Positive criterion | Screen out positive<br>no. | True-positive no.<br>(PPV, %) | True-positive<br>missing no. (%) |
| --- | --- | --- | --- |
| Among TAb(+) Donors in Wuhan (n=590) |  |  |  |
| TAbs-R | 590 | 515 (87.29%) | 0 |
| TAbs/IgM DR | 347 | 339 (97.69%) | 176 (34.17%) |
| TAbs/IgG-RBD DR | 503 | 498 (99.01%) | 17 (2.33%) |
| TAbs/IgG-N DR | 412 | 407 (98.79%) | 108 (20.00%) |
| Among TAb(+) Donors in the other two cities (n=88) |  |  |  |
| TAbs-R | 88 | 4 (4.55%) | 0 |
| TAbs/IgM DR | 17 | 4 (23.53%) | 0 |
| TAbs/IgG-RBD DR | 10 | 2 (20.00%) | 2 (50.00%) |
| TAbs/IgG-N DR | 6 | 0 | 4 (100.00%) |

TAbs, total antibody against SARS-CoV-2; IgG-RBD, IgG antibody against receptor-binding domain (RBD) of the spike protein of SARS-CoV-2; IgG-N, IgG antibody against nucleoprotein of SARS-CoV-2; R, reactive; DR, reactive in double assays; PPV, positive predictive value.

### Potential risk factors associating with asymptomatic infections

As only three confirmed seropositive donors were identified in the cities of Shenzhen and Shijiazhuang, we only performed risk factor analyses among 17,794 donors in Wuhan. As all confirmatory seropositive donors identified in our study declared never had symptoms of COVID-19, they might experience a past asymptomatic infection. Multivariate regression analysis revealed that age and gender were independent risk factors for the presence of antibodies against SARS-CoV-2 (Table 4). Females showed a 1.8-fold (adjusted OR, 95%CI, 1.5 to 2.2, p<0.001) increased risk in comparison with males. Compared with donors with age ranging from 18 to 25, the adjusted OR was 1.1 (95%CI: 0.8 to 1.5, p=0.491) for donors with age of 26 to 35; 1.4 (95%CI: 1.0 to 1.9, p=0.056) for donors with age ranging from 36 to 45; 1.6 (95%CI: 1.2 to 2.3, p=0.010) for donors with age ranging from 46 to 55; and 4.0 (95%CI: 1.9 to 8.6, p=0.002) for donors with an age of 55 or older. No statistical significance was found on donors with different ABO or Rh(D) blood types, ethnicities, occupations, or education levels by single factor logistic regression analysis (p=0.196, 0.764, 0.812, 0.315, and 0.386, respectively). We further analyzed the relationship between antibody titers and age or gender. To avoid the interference from the data of repeated blood donors during the study, we only included samples from 354 donors who only donated once during the study for the analysis. The results revealed that the GMT levels of IgG-N (p=0.001) and ppNAT (p=0.034) showed a significant increased trend with older age, whereas it was not statistically significant for IgG-RBD level (p=0.356). Compared with females, males had comparable GMT levels of IgG-N (p=0.258) and ppNAT (p=0.283) but showed a lower IgG-RBD GMT level (p=0.027).

**Table 4.**
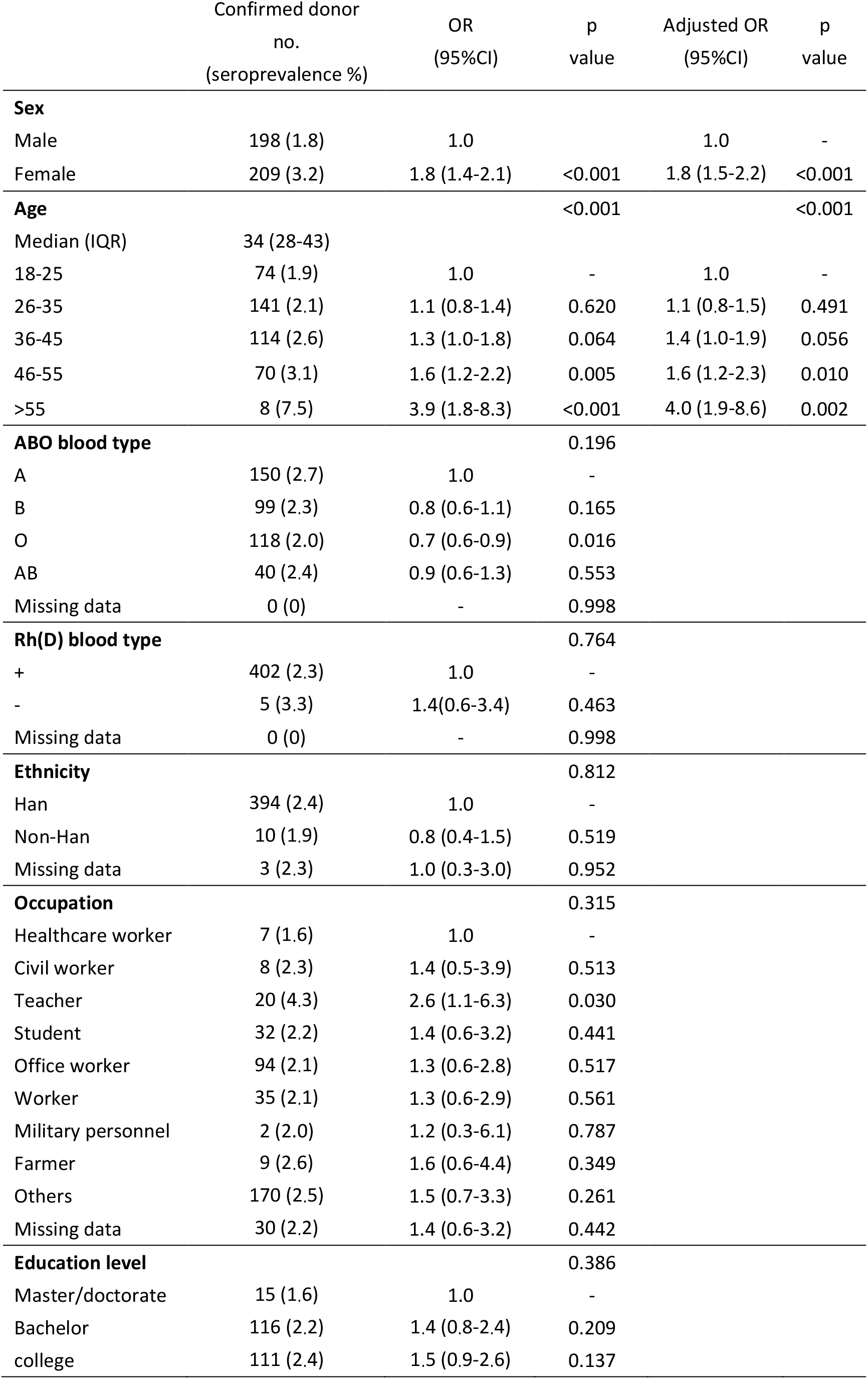
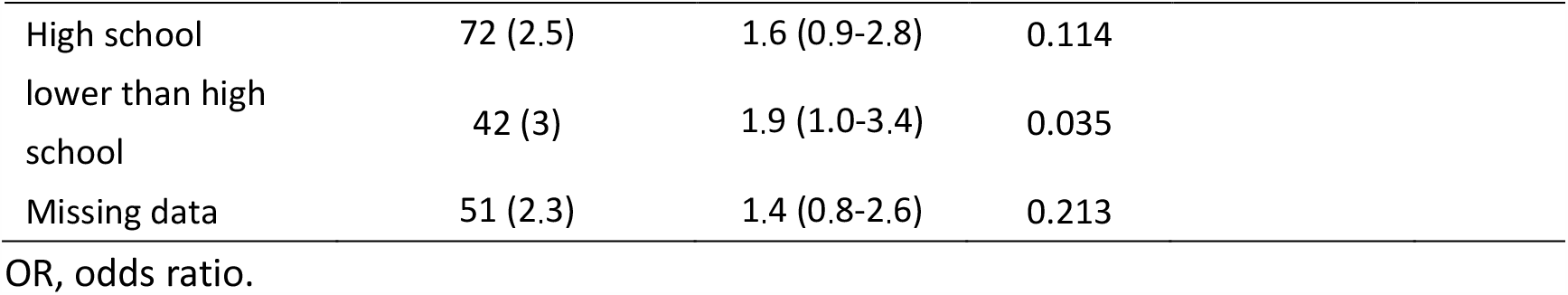
Binary logistic regression analysis of risk factors for the confirmed antibody presence among blood donors in Wuhan

## Discussion

In this study, we systematically investigated the SARS-CoV-2 seroprevalence among large-cohorts of healthy blood donors from different cities in mainland China. Our data clearly demonstrated an overall SARS-CoV-2 seroprevalence of 2.29% (95%CI: 2.08% to 2.52%) in Wuhan, 0.029% (95%CI: 0.0081% to 0.11%) in Shenzhen, and 0.0074% (95%CI: 0.0013% to 0.04%) in Shijiazhuang, respectively. Although Wuhan is the earliest COVID-19 affected city and had most of reported COVID-19 patients in China, the SARS-CoV-2 antibody presence was only noted 2.29% of 17,764 blood donors during January to April 2020, which was lower than that reported in blood donors in Northern France (3%), Netherlands (2.7%), and Italy Milan (5.2%) during the same period.^15,18,19^ Our results suggested that the most of the populations of Wuhan remained uninfected during the early wave of COVID-19. Effective blocked SARS-

CoV-2 spread in China was also evidenced by the extremely low antibody prevalence among donors in Shenzhen and Shijiazhuang. More importantly, in tracking the of antibody presence among Wuhan’s donors in mid to late January, there was no confirmatory positivity among 1614 donors during January 15 to 18. In contrast, only 2 positive donors from 993 donors were found during January 19 to 22 (one donated on January 20, and the other on January 21, Figure 2). Given a TAb seroconversion window of 10-day, the earliest emergence of SARS-CoV-2 in Wuhan’s donors should not earlier than January 10, 2020. The highest seroprevalence (4.63%) was quickly reached at about 2-week after its first emergence (at the 6^th^ week of 2020 during February 2 to 8), and it subsequently slowed to fall during the city quarantine period (January 23 to April 7). After the lockdown easing (during April 8 to 30), we detected a significantly decreased seroprevalence of 2.33% among 10,708 donors (Figure 2). These data provided scientific evidence that demonstrated the effectivity in blocking viral community spread of strict public health measures performed in Wuhan.

Population-based serological surveillance is a critical approach to assess the prevalence of SARS-CoV-2, as well as to estimate herd immunity. Antibody tests with reliable performance are essential both for clinical diagnosis and epidemiological studies. Our study utilized a TAb-based screening strategy followed by a SARS-CoV-2 pseudovirus based neutralization test for final confirmation of the antibody presence (Figure 1). The TAb assay used in this study is a double-sandwich ELISA form, which enables simultaneous detection of IgM, IgG, and IgA against RBD antigen of SARS-CoV-2. Several studies based-on cross-assay comparisons in parallel had demonstrated the TAb-ELISA clearly outperformed all other assays which only detecting single antibody isotype. However, as the reported specificities of TAb-ELISA was ranging from 99.3% to 100%,^20,21^ additional neutralization confirmation should be performed, in particular for reactive samples from RNA-negative asymptomatic individuals. In this study, we determined the neutralization activities of all TAb-positive samples against lentiviral-based pseudotyping SARS-CoV-2 virus as the confirmatory approach. The HIV/lentiviral vectors were widely used in producing of pseudotyping viral particles bearing various highly pathogenic viral envelopes. As all TAb-positive blood donors in our study were HIV-free, it was easy to exclude the possible interfere derived from potential antibodies against surrogate virus proteins, which may present in samples of some uncertain individuals. Our study revealed the PPV value of TAb-based screening was highly dependent on the incidence of confirmed COVID-19 cases among different populations. Referring to neutralization results, the false-positive ratio of TAb-based screening strategy was 0.35% (75/21,483) in Wuhan, 0.36% (25/7040) in Shenzhen, and 0.41% (59/14,271) in Shijiazhuang, without significant difference. However, the PPV values varied from 1.67% (Shijiazhuang) to 87.29% (Wuhan). We noted a TAb/IgG-RBD combining strategy improved the PPV value from 87.29% to 99.01% with little missing (2.33%, 17/515) of the true-positive cases, but its performance was still unsatisfactory in donors of Shenzhen and Shijiazhuang (Table 3). Therefore, neutralization confirmations are indeed required to exclude the false-positive reaction derived from immunoassays that may overestimate the real infection status, in particular for serological studies in a low-prevalence area. In addition, the IgG-RBD titer well correlated with the neutralization titer (Figure 3A), suggested it was a good surrogate for protective immunity assessment when neutralization assay was unavailable.

In COVID-19 patients, gender, age, and ABO blood type were reported to be associated with the occurrence or the development of the disease. Compared with females, male patients have higher mortality (22.2% vs. 10.4%) and require longer hospitalization time.^22^ A study based on 72,314 COVID-19 patients in China found that the gender ratio in Wuhan’s patients was 0.99 (male/female), but the case fatality rate in males was about 1.64-fold higher than that in females,^23^ suggesting an enhanced disease severity for males. Our study revealed that the SARS-CoV-2 seroprevalence was significantly higher among females than males in healthy donors from Wuhan (Adjusted OR: 1.8, 95%CI: 1.5 to 2.2, Table 4). Besides, the average IgG-RBD titer of confirmatory seropositive donors was significantly higher in females than that in males (Figure 4). Taken together, it was possible that females have more probability for asymptomatic infections whereas males are more likely to suffer symptomatic disease. The gender-difference could be attributed to estrogen receptor signaling mediated protections, which had been demonstrated in previous SARS-CoV animal study.^24^ On the other hand, men smoke more than women in China, and smoking may accelerate lung injury and also be associated with a worse clinical outcome of SARS-CoV-2 related disease.^25^ Besides gender, increasing age was also found as an important risk factor for SARS-CoV-2 seropositivity among blood donors (Table 4). Seropositive individuals with older age showed significantly higher titers of IgG-N (Figure 4B) and neutralization antibodies (Figure 4C). The increased risk with age was also noted in both COVID-19 patients and asymptomatic infections, suggesting the older population are more susceptible to SARS-CoV-2 infection than young people.^26,27^ Recent studies suggested that the individuals with blood type A showed higher COVID-19 risk, whereas the blood type O was a possible protective factor. ^28,29^ Our study also noted a lower seroprevalence of SARS-CoV-2 in Wuhan’s donors with blood group O than those with blood type group A in univariate analysis (Table 4). However, the difference was not statistically significant in the logistic regression model. One of the possible reasons is human circulating anti-A antibodies could inhibit the viral adhesion to an ACE2 expressing cell line via blocking the interaction between the virus and its receptor.^29,30^ Overall, female and older-age were the predominant risk factors independently associated with the seropositivity of SARS-CoV-2.

**Figure 4.**
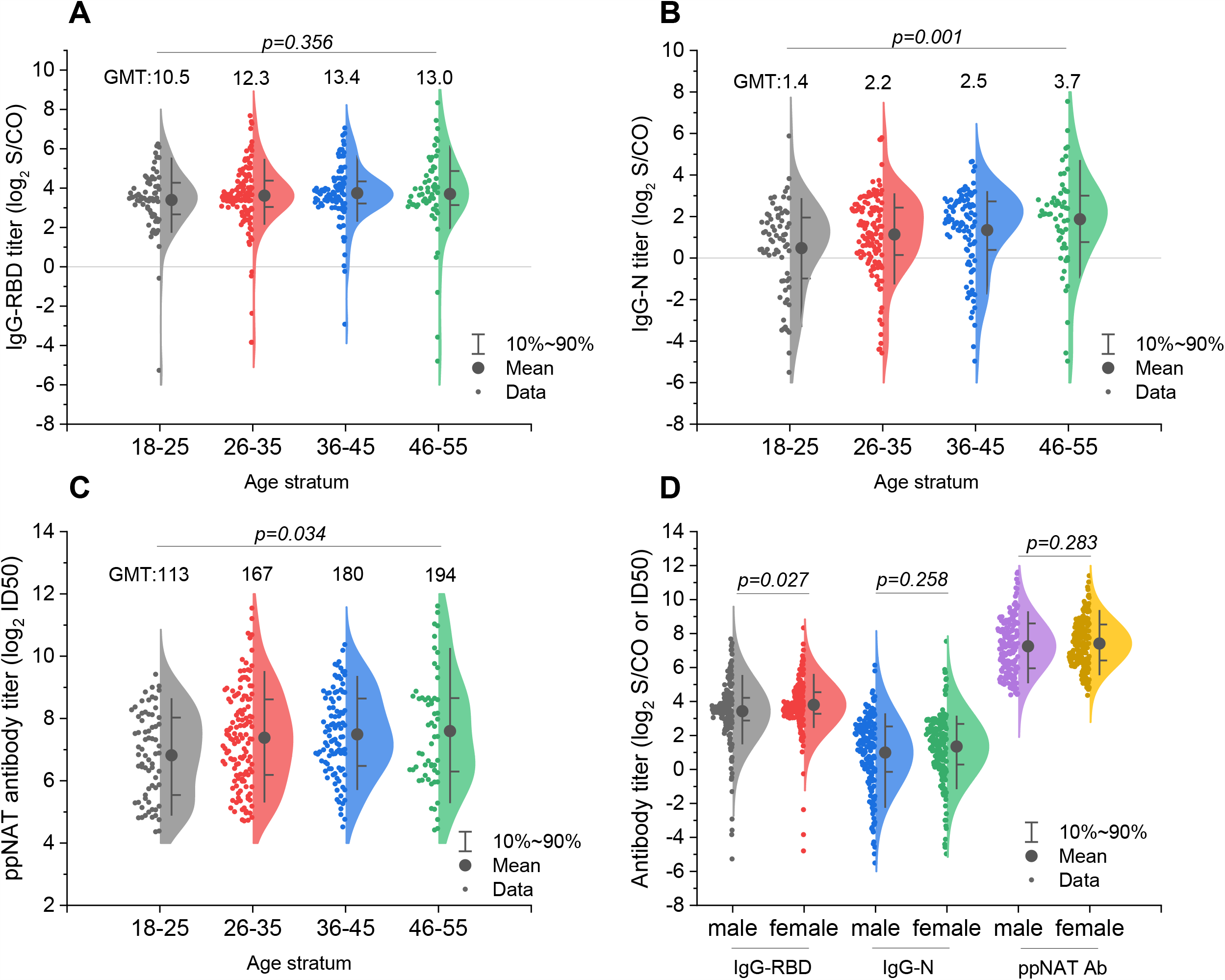
Relationship between specific antibody titer and gender or age. A. Relationship between the titer of IgG-RBD and four different age groups. No statistical significance is shown among different age groups (p=0.356). The horizontal grey line shows the cutoff value of IgG-RBD assay (S/CO=1.0). Geometric mean titers (GMT) of the four groups are 10.6, 12.3, 13.4, and 13.0, respectively. B. Relationship between the titer of IgG-N and four different age groups. A significant difference is shown among different age groups (p=0.001). The horizontal grey line shows the cutoff value of IgG-N assay (S/CO=1.0). Geometric mean titers (GMT) of the four groups are 1.4, 2.2, 2.5, and 3.7, respectively. The IgG-N titer of age group 18-25 is lower than that of group 36-45 (p=0.016) and 46-60 (p=0.001). C. Relationship between the titer of ppNAT ID50 and four different age groups. A significant difference is shown among different age groups (p= 0.034). Geometric mean titers (GMT) of the four groups are 113, 166, 180, and 194, respectively. The ID50 titer of age group 18-25 is lower than that of group 36-45(p=0.035). D. Relationship between the titer of specific IgG or ppNAT ID50 and gender. Female has a higher titer of IgG-RBD antibody (p=0.027), but the titers of IgG-N and ppNAT ID50 antibodies are no difference between male and female (p=0.258 and 0.283).

There are some limitations to our study that should be noted. First, the seroprevalence of our study was derived from data among healthy blood donors, which is a specific population that may have different demographic characteristics to the general population. The “healthy donor effect” should be considered, as people with mild illness or discomfort are not included.^31^ Second, since we didn’t follow all the 410 confirmed positive donors, and have not obtained their detailed information after donation and the nucleic acid testing results of their respiratory tract samples, we couldn’t speculate their possible exposure way and infection status.

In summary, from January to April of 2020, the prevalence of antibody against SARS-CoV-2 among blood donors was 2.29% in Wuhan, 0.029% in Shenzhen, and 0.0074% in Shijiazhuang, which was highly associated with the reported COVID-19 case numbers in these cities. The earliest emergence of SARS-CoV-2 seropositivity among blood donors in Wuhan was identified on January 20, 2020, and the overall seroprevalence in this population showed a downward trend from February to April. Moreover, our study provided a prevalence-dependent antibody testing strategy for population-based serological studies, which highlighted the importance of a confirmatory neutralization test in avoiding the misleading of false-positive results of single immunoassay, in particular for a population in the non-epidemic region.

## Data Availability

No additional data available.

## Contributors

LC, QY and WL conceived the study. LC drafted the first version of the manuscript. HW, ZY, and QY contributed to drafting sections of the manuscript and revised the manuscript. LC, HW, ZY, SY, and QY did data analyses. ZL, WY, WL, XT, WL, WJ, WL, ZJ, XJ, DJ, YR, LY, CW, ZJ, HW, YY, GF did the screening of specific antibodies in labs. HW, ZY, WJ, and MJ did the neutralization tests. ZL, ZJ, HW, CT, ZJ, and XN participated in the study design and helped to draft the manuscript. All authors contributed to the interpretation of data and approved the final manuscript. The corresponding author attests that all listed authors meet authorship criteria and that no others meeting the criteria have been omitted.

## Acknowledgments

All the serological tests against SARS-CoV-2 were provided by Beijing Wantai Biological Pharmacy Enterprise Co.,Ltd. We thank all the staff from Wuhan Blood Center, Shenzhen Blood Center and Hebei Province Blood Center for their hard-working during the pandemic.

## Ethical approval

This study was conformed to the ethical guidelines of the 1975 Declaration of Helsinki and was reviewed and approved by the Medical Ethical Committee of Beijing Hospital (2020BJYYEC-070-01). Written informed consent was obtained from each enrolled donor before donation.

## Data sharing

No additional data available.

## Transparency declaration

The lead authors and manuscript’s guarantors affirms that this manuscript is an honest, accurate, and transparent account of the study being reported; that no important aspects of the study have been omitted; and that any discrepancies from the study as planned have been explained.

## Dissemination to participants and related patient and public communities

The findings of this study will be disseminated to all blood establishments or related institutions caring for individuals tested by specific antibodies against SARS-CoV-2. In addition, our media relation departments will plan to further disseminate through press releases, as well as our institutional websites.

